# Co-infection dynamics of SARS-CoV-2 and respiratory viruses in the 2022/2023 respiratory season in the Netherlands

**DOI:** 10.1101/2024.09.10.24313400

**Authors:** Gesa Carstens, Eva Kozanli, Kirsten Bulsink, Scott McDonald, Mansoer Elahi, Jordy de Bakker, Maarten Schipper, Rianne van Gageldonk-Lafeber, Susan van den Hof, Albert Jan van Hoek, Dirk Eggink

## Abstract

**Objectives:** Evaluation of the presence and effect of SARS-CoV-2 co-infections on disease severity.

**Methods:** We collected both symptom data and nose- and throat samples from symptomatic people during the 2022/2023 respiratory season in a large participatory surveillance study in the Netherlands, and tested these for 18 respiratory viruses including SARS-CoV-2. We compared reported health status, symptoms and odds of having a mono respiratory viral infection or co-infection with SARS-CoV-2 and another respiratory virus.

**Results:** In total 4,655 samples were included with 22% (n=1,017) testing SARS-CoV-2 positive. Of these 11% (n=116) also tested positive for a second respiratory virus. The most frequently occurring co-infections in SARS-CoV-2 positive participants were with rhinovirus (59%; n=69), seasonal coronaviruses (15%; n=17) and adenovirus (7%; n=8). Participants with a co-infection with one of these three viruses did not report more severe disease compared to those with a SARS-CoV-2 mono-infection. The odds of experiencing SARS-CoV-2 co-infection with seasonal coronavirus or rhinovirus were lower compared to the odds of the respective non-SARS-CoV-2 mono-infection (OR: 0.16, CI 95%: 0.10 – 0.24; OR: 0.21 CI 95%: 0.17 – 0.26; respectively).

**Conclusions:** SARS-CoV-2 co-infections with rhinovirus, seasonal coronavirus and adenovirus are frequently observed in the general population, but are not associated with more severe disease compared to SARS-CoV-2 mono-infections. Furthermore, we found indications for inter-virus interaction with rhinovirus and seasonal coronavirus, possibly decreasing risk of co-infection.

**Highlights:** Our study showed viral co-infections in 11% of the SARS-CoV-2 positive participants

SARS-CoV-2 mono-infections led to more severe symptoms than the common cold mono-infections with seasonal coronavirus or rhinovirus

SARS-CoV-2 co-infections with common cold viruses did not lead to worse health status compared to SARS-CoV-2 mono-infections

Odds for mono-infection with common cold virus were higher than for co-infection with SARS-CoV-2 and the respective common cold virus

## INTRODUCTION

The transition of SARS-CoV-2 from pandemic status to endemicity introduced a complex landscape of viral co-circulation, involving not only different SARS-CoV-2 variants but also other respiratory viruses including rhinovirus, influenza virus, adenovirus, respiratory syncytial virus and endemic seasonal coronaviruses. This situation necessitates an examination of prevalence of co-infections, its clinical implications including disease severity, the potential for novel transmission dynamics, and the consequential effects on human susceptibility to a range of viral pathogens.

Co-infections involving SARS-CoV-2 and other respiratory viruses have the potential to modify the clinical manifestation of disease. This makes it more difficult to diagnose and treat these patients. For COVID-19 patients, co-infections are seen in some severe cases (1). In addition, it has been suggested that Influenza A and SARS-CoV-2 co-infections are associated with an increased risk for severe disease and death compared to SARS-CoV-2 mono-infection (2).

Furthermore, this co-circulation could instigate new inter-virus dynamics, with SARS-CoV-2 infection possibly augmenting or diminishing the susceptibility of individuals to other viral infections, thereby influencing their transmission dynamics (3). There are open questions regarding the implications of this co-circulation for the burden of illness, and whether additional public health measures, such as vaccination programs targeting multiple viruses, might be warranted.

To investigate these questions, we conducted a study during the 2022/2023 respiratory season which involved collecting nose and throat swabs by self-sampling from symptomatic individuals in a participatory surveillance cohort in the Netherlands. Samples were tested for SARS-CoV-2 and 18 other respiratory viruses using multiplex PCR. This approach resulted in a comprehensive dataset in which the relationships between SARS-CoV-2 and other co-circulating respiratory viruses could be determined, with particular focus on comparisons between SARS-CoV-2 mono-infection and co-infection with a second respiratory virus in terms of the severity of illness and the set of presented symptoms, as well as an exploratory assessment of susceptibility to co-infection with specific viruses. Specifically, we wished to determine: (i) does SARS-CoV-2 co-infection with another respiratory virus lead to poorer health during the infection (as measured by self-reported health status during the infection episode) compared to the respective mono-infection with another virus or SARS-CoV-2? and (ii) Is there a difference in the odds of infection with other respiratory viruses between participants who tested positive and those who tested negative for SARS-CoV-2?

## METHODS

### Data sources

For this study we used a digital participatory surveillance system *Infectieradar* which was set up in March 2020 in the Netherlands as surveillance system for respiratory diseases independent of health care seeking behavior. A rapid antigen self-test and self-sampling component was added to the platform in the season 2022/2023 enabling pathogen identification in symptomatic or SARS-CoV-2 self-test positive participants. The design has been described in detail previously (4, 5). Briefly, upon registration participants fill in an intake questionnaire collecting sociodemographic and medical background data. Subsequently, participants receive weekly questionnaires asking whether they experienced symptoms consistent with an acute respiratory infection (ARI) and whether they had taken a SARS-CoV-2 self-test. Symptomatic participants are asked to perform a SARS-CoV-2 self-test and to answer additional questions regarding their symptoms, healthcare seeking behavior and health status in the previous week.

A self-sampled nose throat swab (NTS) was requested from a random sample of participants that reported ARI symptoms, i.e. sore throat, cough, runny nose, or dyspnea which had a symptom onset within five days of reporting. The NTS was tested for 18 circulating respiratory viruses.

Moreover, participants who report symptoms consecutively in more than one weekly questionnaire are asked whether the present symptoms belong to the same infection episodes as the previous symptoms. Therefore, participants can report symptoms and their health status more than once per infection episode and each participant can experience several infection episodes throughout the study period.

### Inclusion/exclusion criteria

For this study, we included all participants submitting at least one weekly questionnaire in the respiratory season 2022/2023 (October 1^st^ 2022 to April 30^th^ 2023). Next, swabs were excluded from participants who were not invited and did not have ARI symptoms with onset within 5 days of reporting. For the analysis of the self-sampled swabs, only participants submitting an NTS at least once during the study period were included. Participants could have sent in more than one NTS during the study period, but only one NTS per infection episode.

### Laboratory assays/testing methods

Laboratory methods have been previously described (4). In short, swabs were collected in virus transport medium and RNA extraction was performed using MagNApure 96 (MP96) (Roche). Extracted RNA was subsequently analyzed for the presence of the following respiratory viruses: SARS-CoV-2, influenza A, influenza B, RSV-A, RSV-B, human metapneumovirus, rhino-/enterovirus, adenovirus, parainfluenza-1, parainfluenza-2, parainfluenza-3, parainfluenza-4, bocavirus, seasonal coronavirus NL63, seasonal coronavirus HKU1, seasonal coronavirus OC43, seasonal coronavirus 229E and MERS-CoV, using SARS-CoV-2 specific RT-PCR as described (6) and multiplex real-time PCR (RespiFinder 2SMART, PathoFinder, the Netherlands).. Although the Respifinder cannot officially differentiate between rhino-/enterovirus, additional in-house typing of a representative subset of samples has shown that the majority of these samples are rhinovirus positive. Therefore, in the main text and results we present them as rhinovirus. Samples that test positive for only one pathogen by these assays are considered mono-infections. Bacterial infections or carriage are not considered for this study.

### Primary outcome variable

The main outcome variable was self-reported health status, which was measured in weekly questionnaires. Participants were asked to rate how good or bad their health was in the last week on a scale of 0 to 100, where 100 represented the best imaginable state of health and 0 represented the worst imaginable state of health.

### Exposure variable and covariates

The exposure variable was ‘infection status’ (consisting of: SARS-CoV-2 mono-infection, co-infection SARS-CoV-2 with rhinovirus, co-infection SARS-CoV-2 with adenovirus and co-infection SARS-CoV-2 with seasonal coronaviruses). The following baseline characteristics were included as covariates in multivariable analysis: sex, age-group (<30, 30-39, 40-49, 50-64; 65+ years), education level (none/primary only, middle, higher (i.e., university qualification)), presence of children aged <5 years in the household, presence of children aged 5-18 years in the household (both binary variables), smoker status (non-smoker, ever-smoker), hay fever and/or other allergies and regular use of medication for selected underlying conditions (lung disease (e.g., emphysema, COPD), cardiovascular disease, diabetes; all coded as binary variables).

### Descriptive analysis

We report the observed distribution over the various detected respiratory viruses within our analysis period, and graphically show the temporal characteristics of the detected mono- and co-infections. We describe the distribution of reported symptoms (both ARI symptoms and other possible symptoms), comparing SARS-CoV-2 mono-infection, SARS-CoV-2 co-infection with rhinovirus, co-infection with adenovirus, co-infection with seasonal coronaviruses, and mono-infections with rhinovirus, adenovirus, and seasonal coronaviruses. Reported symptoms from more than one weekly questionnaire per infection episode were pooled.

### Association between participant characteristics and self-reported health status

We performed univariable and multivariable analyses to estimate differences in self-reported health status among all symptomatic individuals within this study population. For this, we used a generalized linear mixed effects model. We incorporated a random intercept for participant ID to account for variation within participants and added a natural spline with three knots to account for time difference between symptom onset and submitting a weekly questionnaire capturing potential variations in health status over the disease course. The multivariable regression model was adjusted for all demographic and comorbidity variables as previously specified.

### Comparison of reported health status between mono- and co-infections with SARS-CoV-2 and respiratory viruses

We used the same generalized linear mixed effects model to estimate the differences in self-reported health status among mono- and coinfected individuals. Based on this, to visually compare the associations of mono-infection and co-infections with SARS-CoV-2 and rhinovirus, adenovirus or seasonal coronaviruses on the health status adjusted for confounders we computed and plotted estimated-marginal means for the health status using the emmeans package (7). Next, we plotted the marginal mean of the health status over time since symptom onset and stratified by age group for the different mono- and co-infections to visually show the progression of the health status during infection.

In an alternative analysis, we analyzed only the lowest reported health status per participant infection episode across mono- and co-infected persons to compare differences in perceived health during the as most severe experienced phase of the infection.

### Exploratory assessment of viral load

To study the possible association between viral load of SARS-CoV-2 mono-infections and co-infections, we performed a linear regression model with Ct value as outcome variable and infection status as independent variable with SARS-CoV-2 mono-infection as reference group. We performed univariate and multivariate analyses, the latter corrected for age group and sex.

### Symptom analysis

We performed multiple multivariable logistic regressions to assess the odds of experiencing various respiratory or constitutional symptoms in case of a non-SARS-CoV-2 mono-infection or SARS-CoV-2 co-infection, using SARS-CoV-2 mono-infection as reference group. The analyses were adjusted for age group and sex.

### Exploratory assessment of co-infection prevalence associated with SARS-CoV-2

We conducted multiple logistic regression analyses for infection with the most common mono-infections; those with rhinovirus or with one of the seasonal coronaviruses. We compared the odds of testing positive for either virus between participants who tested positive and those who tested negative for SARS-CoV-2. The estimated odds ratio (OR) for co-infection with another virus can be interpreted as the association of SARS-CoV-2 infection on susceptibility to that other virus and vice versa. This analysis is necessarily exploratory because other, unmeasured factors could influence the odds of co-infection (e.g., respiratory virus seasonality, and SARS-CoV-2 or influenza vaccination effectiveness).

All analyses were conducted using R statistical software, version 4.3.1 (8).

## RESULTS

### Participants and Sample collection

Within the analysis period (1 Oct 2022 through 30 April 2023), a total of 17,499 participants took part in the *Infectieradar* self-swab study (Fig. S1). Of these, 12,968 (74%) reported ARI symptoms in at least one weekly survey within 5 days of symptom onset, of which 3,498 (27%) reported a positive self-test (Table 1). In total 4,689 NTS samples from 3,945 participants were included in this study and presence of SARS-CoV-2 as well as other respiratory viruses was investigated.

**Table 1.**
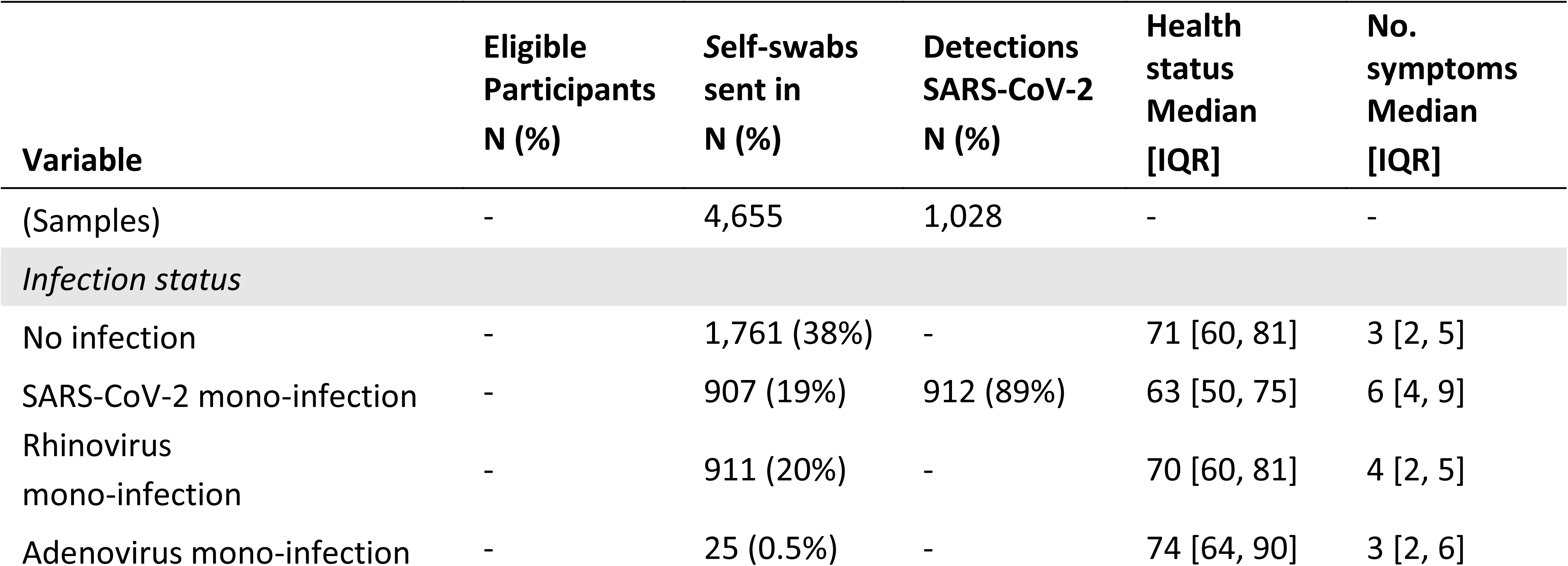
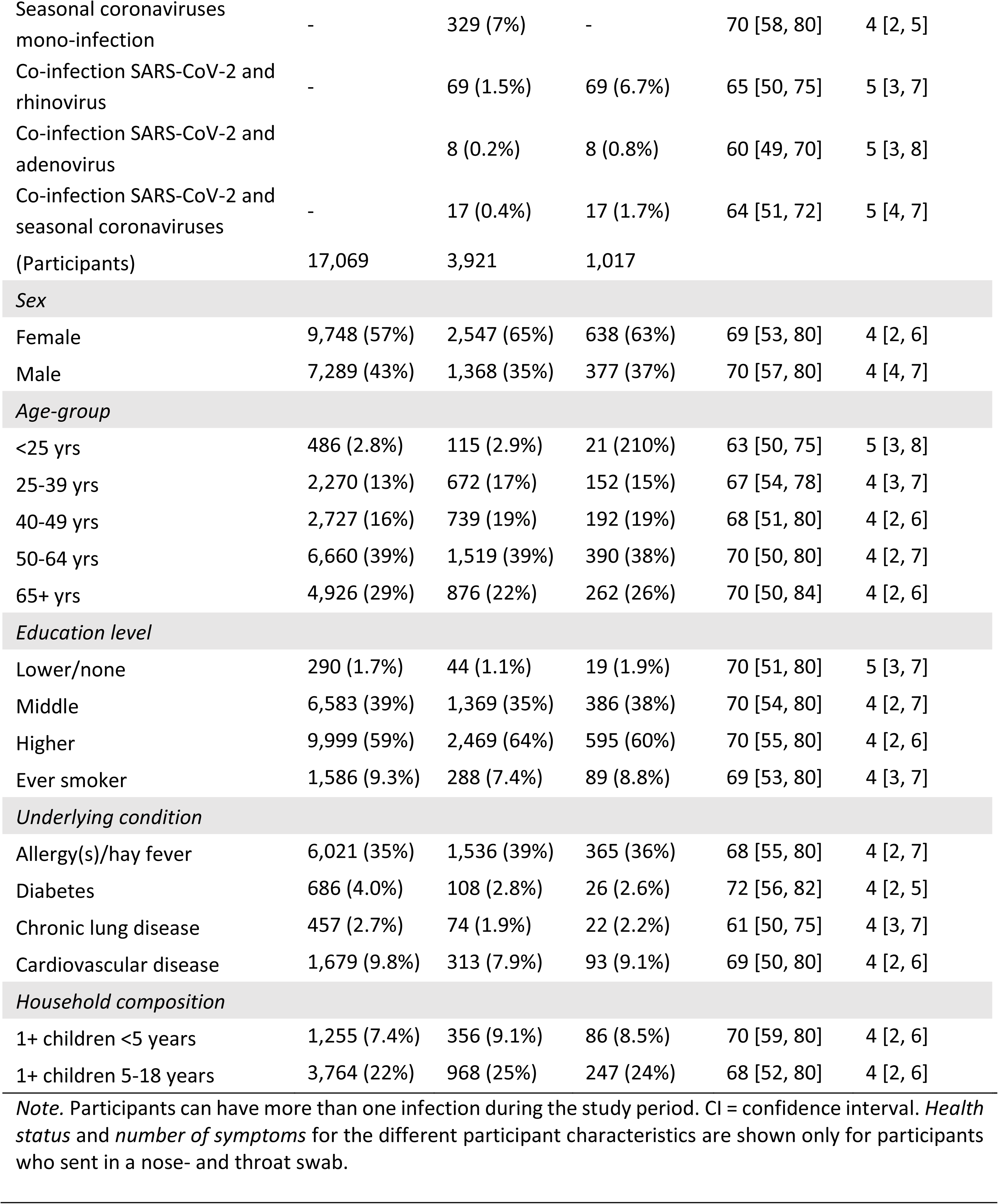
Characteristics of the Infectieradar study population and linked laboratory database.

Our study population had a higher proportion of females (57.2%) than males with the largest age group being 50-64 years (Table 1). The youngest age group (<25) was underrepresented with only 3.3%. The majority of our participants have completed higher education (59%).

### Pathogen detection

1,028 (22%) of samples were SARS-CoV-2 positive measured by SARS-CoV-2 specific RT-PCR. Co-infection with at least one other respiratory virus was detected in 11% (116/1,028) of SARS-CoV-2 positive samples. The most frequently occurring co-infections in SARS-CoV-2 positive participants were rhinovirus (59%; 69/116), seasonal coronaviruses (15%; 17/116), adenovirus (7%, 8/116) or other viruses (Table S1) (20%; 22/116, consisting of influenza A (2/22), influenza B (2/22), bocavirus (2/22), parainfluenza virus (5/22), respiratory syncytial virus (6/22) and human metapneumovirus (5/22)).The corresponding percentages of samples with detected mono-infection were SARS-CoV-2 (19%; 912/4,689), rhinovirus (20%; 923/4,689), adenovirus (0.5%; 25/4,689), and seasonal coronaviruses (7%; 329/4,689).

Co-infections coincided with their respective mono-infection, as shown in Figure 1A and 1B, in which the mono- and co-infections are plotted during the 2022/2023 season.

**Figure 1.**
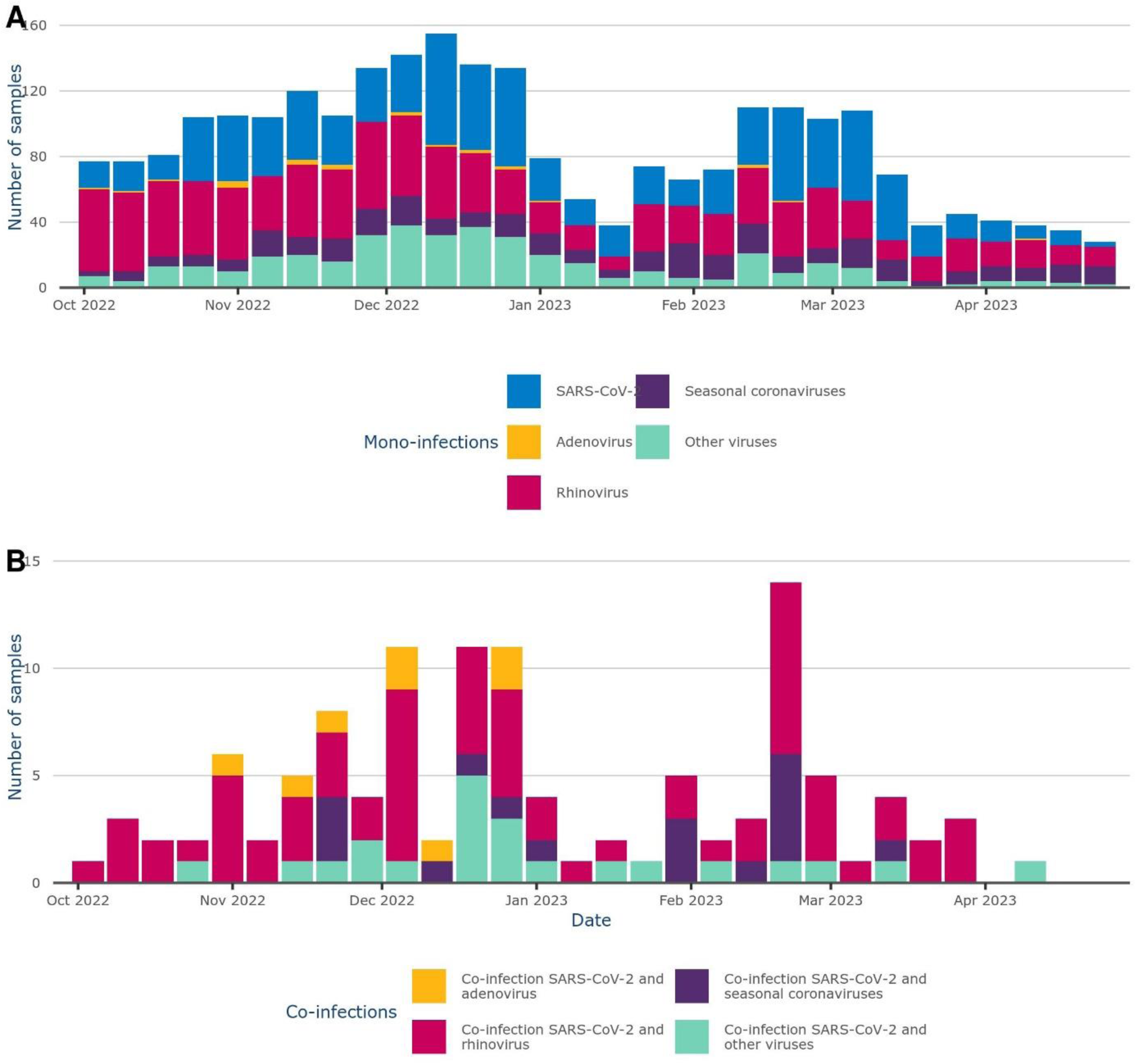
Detected respiratory infections during the 2022/2023 respiratory season in Infectieradar in the Netherlands. Detected mono-infections (A) and co-infections (B) over the analysis period (1 Oct 2022 – 30 April 2023).

### Health status and covariates in the Infectieradar population

Reported health status was analyzed for the symptomatic study population. Most participants reported a health status higher than 50 out of 100 with only slight differences between sex (Fig. S2). We assessed the association between various covariates and self-reported health status by estimating beta coefficients in both univariable and multivariable regression analyses (Fig. 2 and Table S2). The male sex was associated with reporting a higher health status (adjusted beta: 0.17, 95% CI 0.13 to 0.2), as can be seen in Figure 2. Also, age was associated with health status: compared to participants aged 50-64 years, participants of age group <25 and 25-39 were associated with reporting a lower health status (adjusted beta −0.13, 95% CI −0.22 to −0.04 and adjusted beta −0.13, 95% CI −0.18 to −0.08, respectively) while participants of the age group 65+ reported a higher health status (adjusted beta 0.11, 95% CI 0.07 to 0.14). Moreover, participants with allergies and/or hay fever were associated with a lower health status (adjusted beta −0.08, 95% CI −0.11 to −0.05) as well as participants with diabetes (adjusted beta −0.1, 95% CI −0.19 to −0.02) and heart conditions (adjusted beta −0.17, 95% CI −0.23 to −0.12).

**Figure 2:**
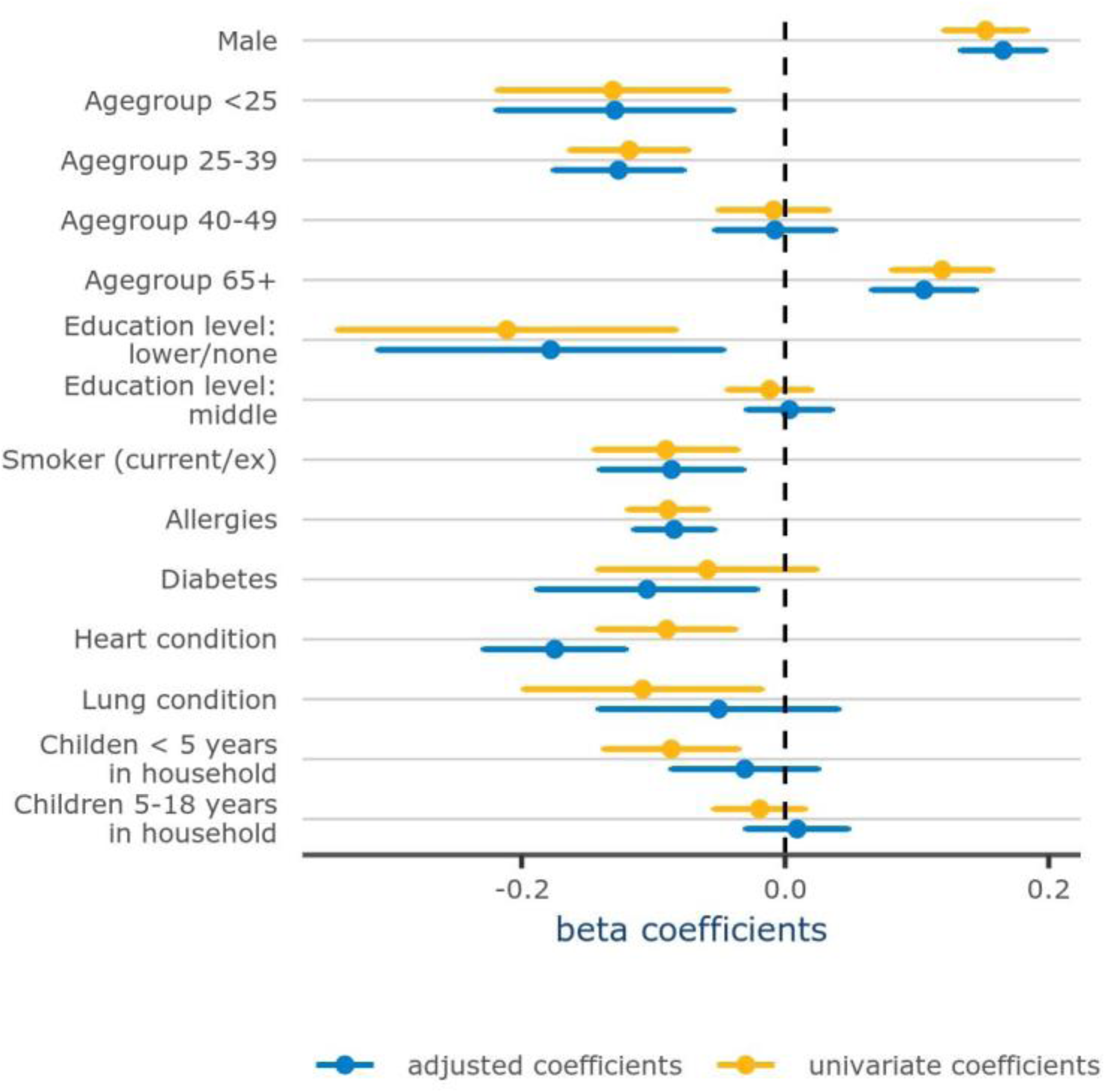
Forest plot of beta coefficients of the linear regression model estimating the association between main participant characteristics with health status during infection. Results of univariable and multivariable regression analyses for all symptomatic participants of our study population with 95% confidence interval.

### Comparison self-reported health status between mono- and co-infected participants

Using the results of all pathogen detections, we estimated the effect of having a mono-infection with SARS-CoV-2, adenovirus, rhinovirus and seasonal coronaviruses and having SARS-CoV-2 co-infections with one of those viruses on perceived health status during infection (Table S3). Figure 3 shows the estimated marginal mean of the health status of each infection. The mean health status was statistically significantly higher for participants with a mono-infection with rhinovirus, seasonal coronaviruses or adenovirus than for those with a SARS-CoV-2 mono-infection, suggesting more severe disease during COVID-19 compared to more traditional respiratory viruses (often referred to as common cold viruses). Interestingly, participants with a co-infection of one of these common cold viruses with SARS-CoV-2 had a similar mean health status compared to participants with a SARS-CoV-2 mono-infection.

**Figure 3:**
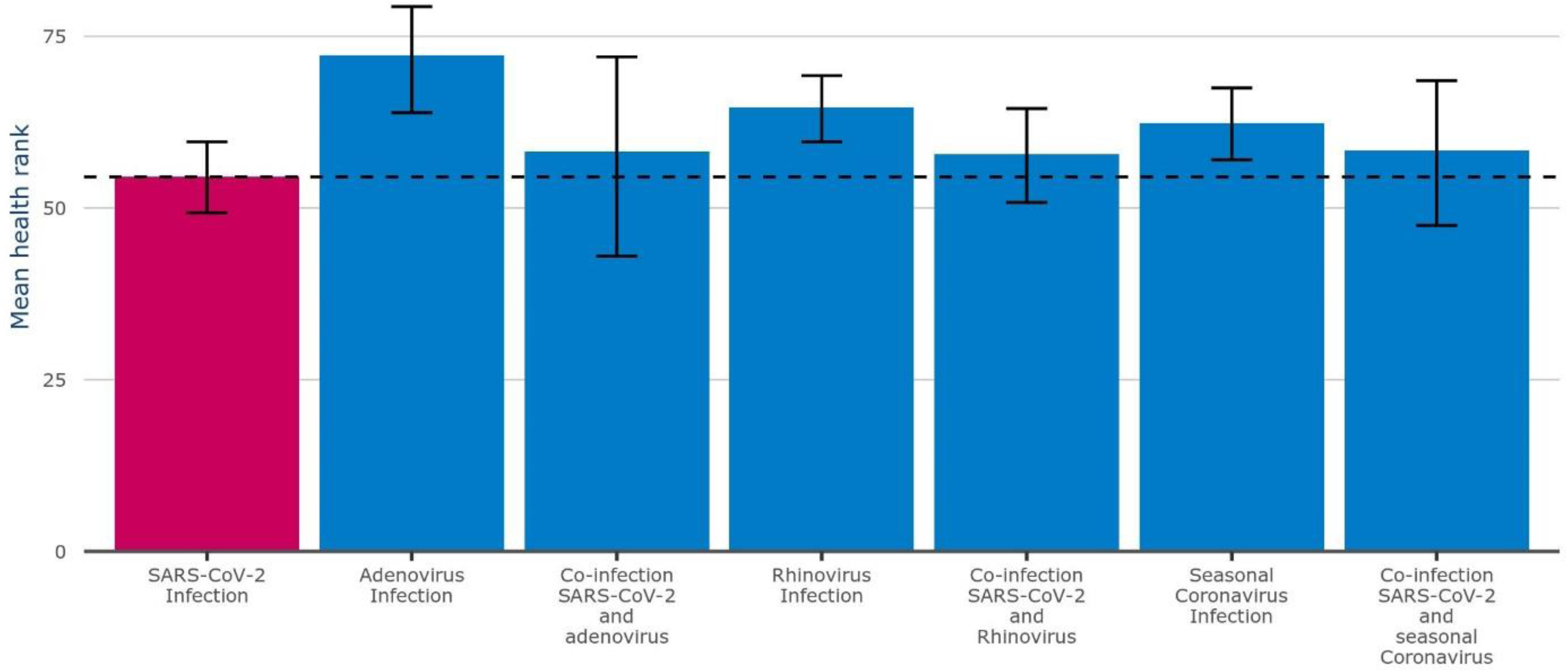
Mean health status during infection. Estimated marginal means of health status and 95% Confidence level per infection. Horizontal line indicating the estimated-marginal mean health status for participants with SARS-CoV-2 mono-infection infection.

Additionally, we observed that the estimated marginal mean health status varies over time depending on the day since symptom onset (Fig. S3). Across all age groups and infections, the lowest health is reported around day 4-6 since symptom onset before gradually increasing again, with older age groups consistently reporting a higher health status during the whole infection episode compared to the lowest age group. Due to the change in health status during disease progression we adjusted the main analysis and performed an alternative analysis.

In this alternative analysis, we investigated the lowest reported health status per infection episode instead of the mean health status in the model to compare severity during the most severe phase of different infections as perceived by the participants (n = 4,950) instead of the health status over the complete episode. All trends determined in the main analysis were similar in this analysis (Fig. 4 and Table S4). Overall, we found SARS-COV-2 cases more severe disease than traditional common cold viruses, however SARS-CoV-2 co-infections do not cause worse health status compared to SARS-CoV-2 mono-infections.

**Figure 4.**
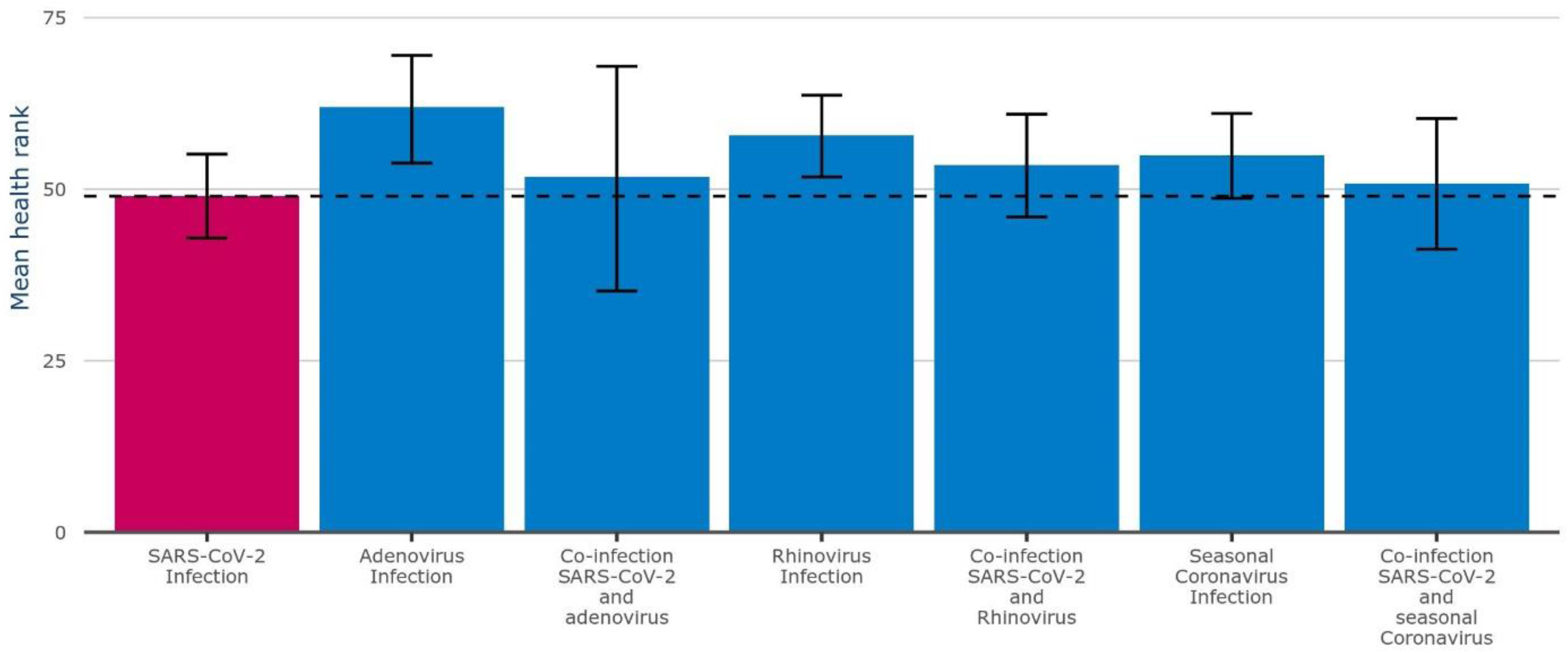
Results of the alternative analysis using only the lowest health status per infection period as model input. Estimated marginal means of the health status with 95% Confidence level per infection. Horizontal line indicating the estimated-marginal mean health status for participants with SARS-CoV-2 mono-infection infection.

As no difference in disease severity was observed between SARS-CoV-2 mono-infections and co-infections, it is interesting to investigate the effect of a co-infection on SARS-CoV-2 viral load. Viral load has been related to disease severity before (9). And in addition, viral load might influence the risk of co-infection. We therefore compared Ct values as a proxy of viral load between SARS-CoV-2 mono-infections and co-infections. Mono-infections of SARS-CoV-2 are associated with a significantly lower Ct value (p value <0.01), reflecting higher viral load, as compared to co-infections with rhinovirus and SARS-CoV-2 (mean Ct value: 22.3 and 24.0, respectively, Table S5). We could not confirm higher Ct values for SARS-CoV-2 co-infections with adenovirus and coronaviruses (mean Ct value: 21.6 and 24.3, respectively), likely caused by a lower number of cases.

### Difference in symptoms between mono- and co-infected individuals

The median number of reported different symptoms of participants with SARS-CoV-2 mono-infection was 6 [Interquartile range (IQR): 4-9], which was higher than reported by participants with other mono-infections (rhinovirus: 4 [IQR 2-5]; adenovirus: 3 [IQR 2-6]; seasonal coronaviruses: 4 [IQR 2-5]) and co-infections (co-infection SARS-CoV-2 with rhinovirus: 5 [IQR 3-7]; co-infection SARS-CoV-2 with adenovirus: 5 [IQR 3-8]; co-infection SARS-CoV-2 with seasonal coronaviruses: 5 [IQR 4-7], Table 1). In line with this, 40% (n = 365) of the participants with SARS-CoV-2 mono-infection took sick leave, while sick leave was significantly less common among participants with other mono-infections (11% (n = 97), p-value <0.001 for rhinovirus; 16% (n = 4), p-value 0.01 for adenovirus; and 10% (n = 32), p-value < 0.001 for seasonal coronavirus; Table S6). However, SARS-CoV-2 co-infected participants took sick leave significantly more often than their mono-infected non-SARS-CoV-2 counterparts but not more than participants with SARS-CoV-2 mono-infection (rhinovirus co-infection: 33% (n = 23), p-value 0.31; adenovirus co-infection: 63% (n = 5), p-value 0.28; seasonal coronavirus co-infection: 35% (n = 6), p-value 0.81).

Of the participants mono-infected with SARS-CoV-2, 87% reported rhinitis, as well as all participants co-infected with adenovirus (100%) and 94% of participants co-infected with seasonal coronaviruses (Fig. 5). Cough, a sore throat and sneezing were also reported frequently by most participants. Compared to SARS-CoV-2 mono-infection, the odds of experiencing respiratory or constitutional symptoms were mainly similar for SARS-CoV-2 co-infected participants (Fig. S4). However, SARS-CoV-2 and rhinovirus co-infected individuals had slightly lower odds of experiencing headache, sneeze, pain and chills.

**Figure 5:**
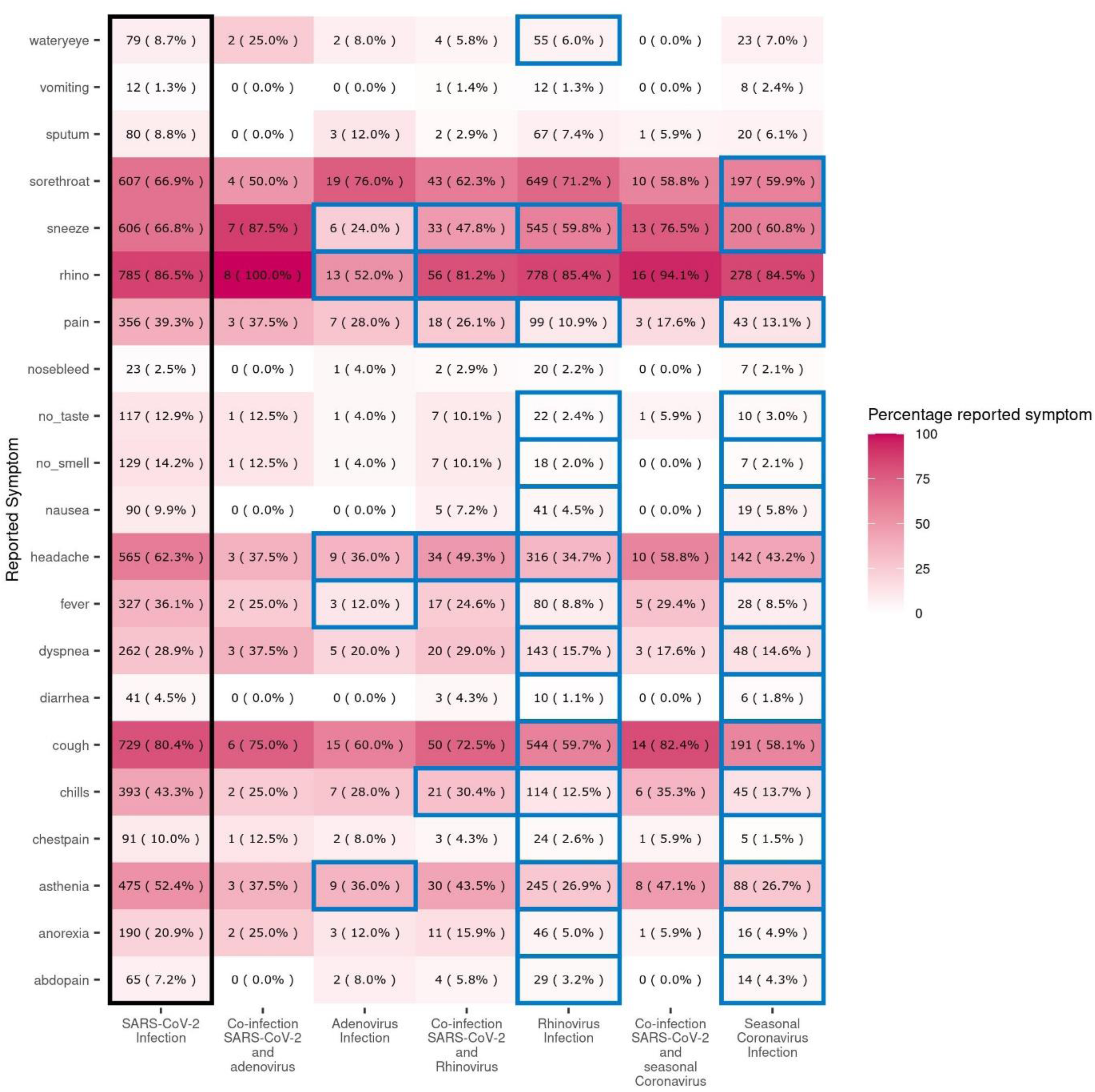
Overview of symptom prevalence. Percentage reported symptoms per infection (SARS-CoV-2 mono-infection, SARS-CoV-2 co-infection with adenovirus, co-infection with rhinovirus, co-infection with seasonal coronaviruses, and mono-infection with rhinovirus, adenovirus, and seasonal coronaviruses) among all participants with lab samples (*n*=3,963). Numbers in the heatmap correspond to N (%). Colored boxes indicate symptoms that are significantly less reported (blue boxes) compared to the reference group SARS-CoV-2 mono-infection (black box). There were no symptoms which were significantly more often reported compared to the reference group.

### Exploratory assessment of co-infection prevalence associated with SARS-CoV-2

Next, we investigated whether infection with SARS-CoV-2 did alter the odds of infection with another respiratory virus (Fig. 6). Odds for co-infection with SARS-CoV-2 combined with either seasonal coronavirus or rhinovirus are lower than mono-infection with one of these non-SARS-CoV-2 viruses (OR: 0.16, CI 95%: 0,10 to 0.24, n = 17; OR: 0.21, CI 95%: 0.17 to 0.24, n = 69; respectively). The number of adenovirus co-infections was too low to perform this analysis on.

**Figure 6:**
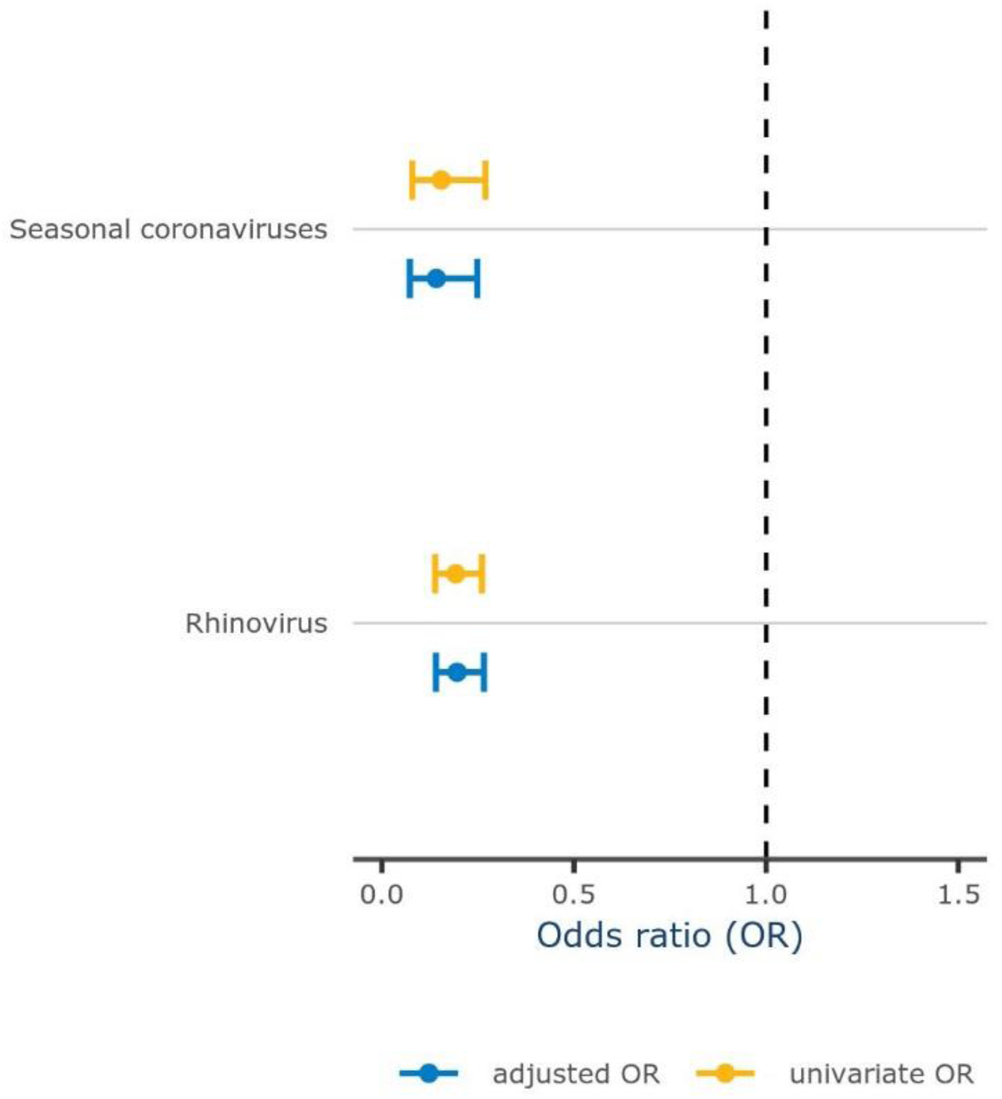
Odds ratio for co-infection with SARS-CoV-2 and other pathogens compared to SARS-CoV-2 mono-infection. The plot shows odds ratios for co-infection with seasonal coronavirus (n=17) and rhinovirus (n=69).

## DISCUSSION

We examined the associations between respiratory mono-infections and co-infections with SARS-CoV-2, and reported symptoms and perceived health in the Netherlands during the transition of SARS-CoV-2 from a pandemic to an endemic virus. Co-infection of SARS-CoV-2 with another respiratory virus was found in 11% of SARS-CoV-2 positive participants; these co-infections included mainly rhinovirus, seasonal coronaviruses and adenovirus. Compared with SARS-CoV-2 mono-infection, participants’ self-reported health status for a mono-infection with these three common cold viruses was higher, suggesting less severe disease. However, a co-infection of SARS-CoV-2 with these three common cold viruses resulted in a similar health status compared to SARS-CoV-2 mono-infection. Across all mono- and co-infections similar symptoms were reported with rhinitis, cough, sore throat and sneezing being the most frequent symptoms. However, SARS-CoV-2 and rhinovirus co-infected individuals had lower odds of experiencing headache, sneeze, pain and chills than individuals infected only with SARS-CoV-2.

A meta-analysis of studies reporting co-infection of SARS-CoV-2 with other respiratory viruses up to August 2021 found a pooled co-infection prevalence of 5.01% (95% CI 3.34%–7.27%) among those with SARS-CoV-2 (10). This is lower than the 11% we found in our study. However, our study period took place between October 2022 and May 2023. During this period, there was a higher circulation of SARS-CoV-2 (with omicron variants) (11) compared to the period before August 2021. Also, there were fewer nonpharmaceutical interventions such as facemasks and social distancing measures in place, leading to heightened circulation of all respiratory viruses and thus to a higher probability of (co-)infection (12). This highlights the impact of temporal fluctuations and co-circulation of respiratory viruses on co-infection rates, which may vary across different periods and geographic locations.

Interestingly, we observed low numbers of co-infections with influenza. Unlike the findings from a study in Missouri, USA conducted between October 2021 and January 2022, in which a prevalence of 33% was reported for influenza co-infection among persons with SARS-CoV-2 (13). Even though in the Netherlands the 2022/2023 influenza season lasted until approximately week 11 (i.e., 13-19 March 2023) (11) we detected 110 Influenza mono-infections and only 4 influenza and SARS-CoV-2 co-infections (Table S1).

There is limited literature on SARS-CoV-2 co-infections among those who do not seek health care. A meta-analysis, in which the vast majority of included studies had been conducted among adult-patients hospitalized for SARS-CoV-2, found an increased odds of mortality (OR=2.84; 95% CI: 1.42–5.66) associated with bacterial, fungal or viral co-infection (14). This increase in severity in SARS-CoV-2 co-infected patients compared to mono-infected patients is not confirmed with our results. We observed that co-infected individuals reported a similarly perceived health status compared to participants mono-infected with SARS-CoV-2, with a point estimate which indicates slightly less severe disease. However, we only included co-infections with respiratory common cold viruses, and did not test for many bacteria and fungi. Therefore, some of those labeled as SARS-CoV-2 mono-infections could have been co-infected with one of those untested pathogens. Additionally, we observed that viral loads were lower in co-infected individuals, which could have influenced why the health status was not more severe for SARS-CoV-2 co-infections compared to SARS-CoV-2 mono-infections. Furthermore, these lowered viral loads in the co-infection group could be an indication for inter-virus interaction.

In our exploratory analysis, we observed lower odds of experiencing SARS-CoV-2 co-infections with both seasonal coronaviruses and rhinovirus compared to experiencing mono-infection with those non-SARS-CoV-2 viruses. Seasonality of viruses might affect how much co-circulation of different respiratory viruses occurs and therefore also affects the number of co-infections with these viruses and SARS-CoV-2. A lack of co-circulation or co-infection between respiratory viruses can suggest interaction, i.e. competition for susceptible hosts between those viruses or internal seasonality of those viruses (15). During our study period in the respiratory season (October to May) we observed co-circulation of the compared respiratory viruses with SARS-CoV-2 indicating the possibility of co-infections. However, our exploratory assessment of the odds of SARS-CoV-2 co-infection indicates that we observed fewer cases than expected based on the prevalence of the non-SARS-CoV-2 respiratory viruses. This finding could suggest virus-virus interplay between SARS-CoV-2 and rhinovirus as well as seasonal coronaviruses. For other pathogens numbers were too low to assess this in this study.

However, without knowing the timing of infection of the viruses in the host and the subsequent immune response, it is difficult to understand the underlying mechanisms that influence SARS-CoV-2 interplay with other respiratory viruses. Viral interference is often proposed as a cause for this interplay in which case presence of one virus could inhibit entrance or replication of the other virus through mechanisms of the virus itself, or of its host. Cellular overlap and for example use of similar receptors for cell entry could affect co-infection rates (16, 17). Furthermore, it was previously suggested that as host innate immunity, including interferon response, increases upon the first infection, susceptibility for a second infection is altered (18). Previous studies have shown that the presence of rhinovirus in the respiratory endothelium can block SARS-CoV-2 viral replication (19), while SARS-CoV-2 primary infection was shown to not impact rhinovirus replication (20). Evidence for the interplay between seasonal coronaviruses and SARS-CoV-2 is less clear: several studies have suggested that cross-immunity exists for SARS-CoV-2 and seasonal coronaviruses (17, 21), while other studies found increased SARS-CoV-2 infection susceptibility and increased disease severity associated with pre-existing immunity for seasonal coronaviruses (22). Although these interplays are likely to explain the reduced odds of SARS-CoV-2 co-infection with seasonal coronaviruses or rhinovirus, a simplistic model can hardly capture these dynamics. More intricate models require more information on the timing and dynamics of infections and host responses to define virus interactions. Adding to the theory of viral interference, we observed that co-infections of SARS-CoV-2 and rhinovirus have lower viral load than SARS-CoV-2 mono-infection. This could mean that the presence of one virus could obstruct the susceptibility or efficient replication of another virus. However, these findings should be further investigated to confirm viral interference. To further explore this, *in vitro* studies might be more appropriate or cohort studies using dense sampling allowing detection of viral dynamics, including low viral loads of co-infections. Additionally, future work using sequential sampling could focus on the implications of the order of infection on the susceptibility to acquire co-infection.

In this study, we found that less people called sick with a mono-infection compared to those with a co-infection of these same viruses with SARS-CoV-2. This observation does not support an effect of behavior, in the form of isolation, on co-infection susceptibility, as mono-infected individuals are not more isolated than co-infected individuals. Moreover, we observed more sick leave for SARS-CoV-2 mono-infections as compared to other mono-infections. This suggests that SARS-CoV-2 infected individuals may have experienced more severe disease, as also indicated by the reported health status. However, as people had a self-test result, previous advice to stay at home when being infected, likely also affected behavior.

The main strength of this study is the community-based population consisting of voluntary participants from the *Infectieradar* surveillance system who self-tested for SARS-CoV-2 and present with mild symptoms. Due to the weekly questionnaires and self-sampling, we were able to catch infections in an early phase of the disease and collect information on perceived health and symptoms during the infection. Potential limitations concern representativeness: compared with the general population, *Infectieradar* participants are more likely to be female, ages 50-64 years are over-represented, and are more highly educated and have lower prevalence of underlying health problems (5). Moreover, we cannot estimate the role of behavior in co-infection prevalence. Self-reporting introduces biases in perceived health status and could lead to missing cases because of more severe diseases being not reported in *Infectieradar*. Furthermore, studying individuals with ARI symptoms can introduce selection bias, as viral status and viral load may affect the probability of symptoms and consequently the likelihood of individuals being included in the study population (23). Therefore, our observed odds ratios of SARS-CoV-2 co-infections might not reflect the full population-level interactions between SARS-CoV-2 and other respiratory viruses due to unknown selection probabilities.

In conclusion, we show that, within a cohort of the general population in the Netherlands in the respiratory season of 2022 to 2023, SARS-CoV-2 co-infections with rhinovirus, seasonal coronavirus and adenovirus did not lead to a worse perceived disease severity compared to SARS-CoV-2 mono-infections. In our study the odds of experiencing SARS-CoV-2 co-infection with rhinovirus and seasonal coronaviruses were lower than for experiencing the respective non-SARS-CoV-2 mono-infections, suggesting a form of viral interference.

## Data Availability

The processed data required to reproduce the findings of this study are available from the corresponding author upon reasonable request. Due to confidentiality concerns, access to the raw data may be restricted.

## Ethics

The samples were collected as part of *Infectieradar*. The protocol for *Infectieradar* was approved by the Medical Ethics Review Committee Utrecht (reference number: WAG/avd/20/008757; protocol 20-131) was obtained given the nature of data collection. All participants provided informed consent upon registration, which included agreement to the privacy statement and described the processing of personal data and research results, website security measures taken, and how to file a complaint. Participants were eligible to withdraw from the study at any time. Individuals had to be 16 years or over to be able to participate.

## Acknowledgements

We thank the participants of Infectieradar for their weekly contributions and submission of samples. Without their contribution this project won’t have been a success. Furthermore, we thank our colleagues who help to maintain and run the Infectieradar-cohort and handling the samples. Developing and running this study has been a huge team effort.

