## Supplementary Material for "Co-infection dynamics of SARS-CoV-2 and respiratory viruses in the 2022/2023 respiratory season in the Netherlands"

### SUPPLEMENTARY TABLES

**Table S1. All other detected mono-infections and low frequency co-infections with SARS-CoV-2.** This table shows all co-infections that were observed in our study at low frequency and other mono-infections that were observed

| Pathogen | Number of positive samples | |
|---|---|---|
| | *Mono-infections* | *Co-infections with SARS-CoV-2* |
| Bocavirus | 3 | 2 |
| Human metapneumovirus | 119 | 5 |
| Influenza A | 50 | 2 |
| Influenza B | 60 | 2 |
| Parainfluenza viruses | 67 | 5 |
| Respiratory syncytial virus | 109 | 6 |

**Table S2. Overview of model output of the generalized linear mixed effect model estimating the association between participant characteristics and perceived health status among symptomatic participants**

| | Univariate | | | Multivariate | | |
|---|---|---|---|---|---|---|
| **Variable** | **beta** | **95% CI** | **P value** | **beta** | **95% CI** | **P value** |
| *Sex* | | | | | | |
| Female | Ref. | | | | | |
| Male | 0.15 | 0.12 to 0.18 | <0.001 | 0.17 | 0.13 to 0.2 | <0.001 |
| *Age* | | | | | | |
| <25 | -0.13 | -0.22 to 0.04 | <0.001 | -0.13 | -0.22 to -0.04 | <0.001 |
| 25-39 | -0.12 | -0.16 to -0.07 | <0.001 | -0.13 | -0.18 to -0.08 | <0.001 |
| 40-49 | -0.01 | -0.05 to 0.03 | 0.68 | -0.01 | -0.05 to 0.04 | 0.74 |
| 50-64 | Ref. | | | | | |
| 65+ | 0.12 | 0.08 to 0.16 | <0.001 | 0.11 | 0.07 to 0.14 | <0.001 |
| *Education level* | | | | | | |
| higher | Ref. | | | | | |
| lower/none | -0.21 | -0.34 to -0.08 | <0.001 | -0.18 | -0.31 to -0.05 | 0.01 |
| middle | -0.01 | -0.04 to 0.02 | 0.47 | 0.01 | -0.03 to 0.04 | 0.87 |
| Smoker | -0.09 | -0.14 to -0.04 | <0.001 | -0.09 | -0.14 to -0.03 | <0.001 |
| *Co-morbidities* | | | | | | |
| Allergies | -0.09 | -0.12 to -0.06 | <0.001 | -0.08 | -0.11 to -0.05 | <0.001 |
| Diabetes | -0.06 | -0.14 to 0.02 | 0.16 | -0.1 | -0.19 to -0.02 | 0.01 |
| Chronic lung disease | -0.11 | -0.2 to -0.02 | 0.02 | -0.05 | -0.14 to 0.04 | 0.28 |
| Cardiovascular disease | -0.09 | -0.14 to -0.04 | <0.001 | -0.17 | -0.23 to -0.12 | <0.001 |
| *Household composition* | | | | | | |
| 1+ children < 5 years | -0.09 | -0.14 to -0.04 | <0.001 | -0.03 | -0.09 to 0.03 | 0.28 |
| 1+ children 5-18 years | -0.02 | -0.05 to 0.02 | 0.27 | 0.01 | -0.03 to 0.05 | 0.65 |

**Table S3 Overview of model output of the generalized linear mixed effect model estimating the association between infection and perceived health status among participants with NTS.** Model is corrected for sex, age group, education level, co-morbidities and household composition

| | Univariate | | | Multivariate | | |
|---|---|---|---|---|---|---|
| **Variable** | **beta** | **95% CI** | **P value** | **beta** | **95% CI** | **P value** |
| *Infection* | | | | | | |
| SARS-CoV-2 mono-infection | Ref. | | | Ref. | | |
| Rhinovirus mono-infection | 0.38 | 0.3 to 0.46 | <0.001 | 0.42 | 0.34 to 0.5 | <0.001 |
| Adenovirus mono-infection | 0.65 | 0.32 to 0.99 | <0.001 | 0.78 | 0.44 to 1.12 | <0.001 |
| Seasonal coronaviruses mono-infection | 0.27 | 0.17 to 0.37 | <0.001 | 0.33 | 0.22 to 0.43 | <0.001 |
| Co-infection SARS-CoV-2 and rhinovirus | 0.1 | -0.11 to 0.3 | 0.36 | 0.13 | -0.07 to 0.34 | 0.21 |
| Co-infection SARS-CoV-2 and adenovirus | 0.07 | -0.52 to 0.65 | 0.82 | 0.15 | -0.44 to 0.74 | 0.62 |
| Co-infection SARS-CoV-2 and seasonal coronaviruses | 0.16 | -0.24 to 0.55 | 0.44 | 0.16 | -0.24 to 0.55 | 0.44 |

**Table S4 Alternative analysis.** Model output of the generalized linear mixed model estimating the association between infection and perceived health status among participants with NTS using only the lowest reported health status per infection episode. Model is corrected for sex, age group, education level, co-morbidities and household composition.

| | Univariate | | | Multivariate | | |
|---|---|---|---|---|---|---|
| **Variable** | **beta** | **95% CI** | **P value** | **beta** | **95% CI** | **P value** |
| *Infection* | | | | | | |
| SARS-CoV-2 mono-infection | Ref. | | | Ref. | | |
| Rhinovirus mono-infection | 0.34 | 0.28 to 0.4 | <0.001 | 0.36 | 0.3 to 0.42 | <0.001 |
| Adenovirus mono-infection | 0.54 | 0.3 to 0.78 | <0.001 | 0.53 | 0.29 to 0.77 | <0.001 |
| Seasonal coronaviruses mono-infection | 0.24 | 0.16 to 0.32 | <0.001 | 0.24 | 0.16 to 0.32 | <0.001 |
| Co-infection SARS-CoV-2 and rhinovirus | 0.15 | -0.05 to 0.35 | 0.8 | 0.18 | -0.02 to 0.38 | 0.07 |
| Co-infection SARS-CoV-2 and adenovirus | 0.09 | -0.59 to 0.76 | 0.15 | 0.11 | -0.54 to 0.76 | 0.74 |
| Co-infection SARS-CoV-2 and seasonal coronaviruses | -0.04 | -0.36 to 0.26 | 0.75 | 0.07 | -0.23 to 0.38 | 0.64 |

**Table S5 Univariate and multivariate SARS-CoV-2 Ct value comparison of participants with mono- and co-infection**

| Variable | Mean Ct value | Univariate P value | Multivariate P value |
|---|---|---|---|
| Mono-infection SARS-CoV-2 | 22.3 | Ref. | Ref. |
| Co-infection SARS-CoV-2 and rhinovirus | 24.6 | **<0.01** | **<0.01** |
| Co-infection SARS-CoV-2 and adenovirus | 21.6 | 0.71 | 0.59 |
| Co-infection SARS-CoV-2 and seasonal coronaviruses | 24.3 | 0.07 | 0.06 |

**Table S6 Differences in sick-leave between different infections.**

| | Sick-leave | | | |
|---|---|---|---|---|
| | Yes, N (%) | No, N (%) | p-value[1] | p-value[2] |
| SARS-CoV-2 mono-infection | 365 (40%) | 542 (60%) | Ref. | - |
| Rhinovirus mono-infection | 97 (11 %) | 814 (89%) | <0.001 | Ref. |
| Co-infection SARS-CoV-2 and rhinovirus | 23 (33%) | 46 (67%) | 0.31 | <0.001 |
| Adenovirus mono-infection | 4 (16 %) | 21 (84%) | 0.01 | Ref. |
| Co-infection SARS-CoV-2 and adenovirus | 5 (63%) | 3 (38%) | 0.28 | 0.02 |
| Seasonal coronaviruses mono-infection | 32 (0.7 %) | 297 (90 %) | <0.001 | Ref. |
| Co-infection SARS-CoV-2 and seasonal coronaviruses | 6 (35%) | 11 (65%) | 0.81 | 0.01 |

[1]p-values indicate pairwise comparisons between participants with SARS-CoV-2 mono infection and other infections. Fisher exact test

[2]p-values indicate pairwise comparison of participants with a SARS-CoV-2 co-infection with another respiratory virus and mono-infection with the respective respiratory virus. Fisher exact test

### SUPPLEMENTARY FIGURES

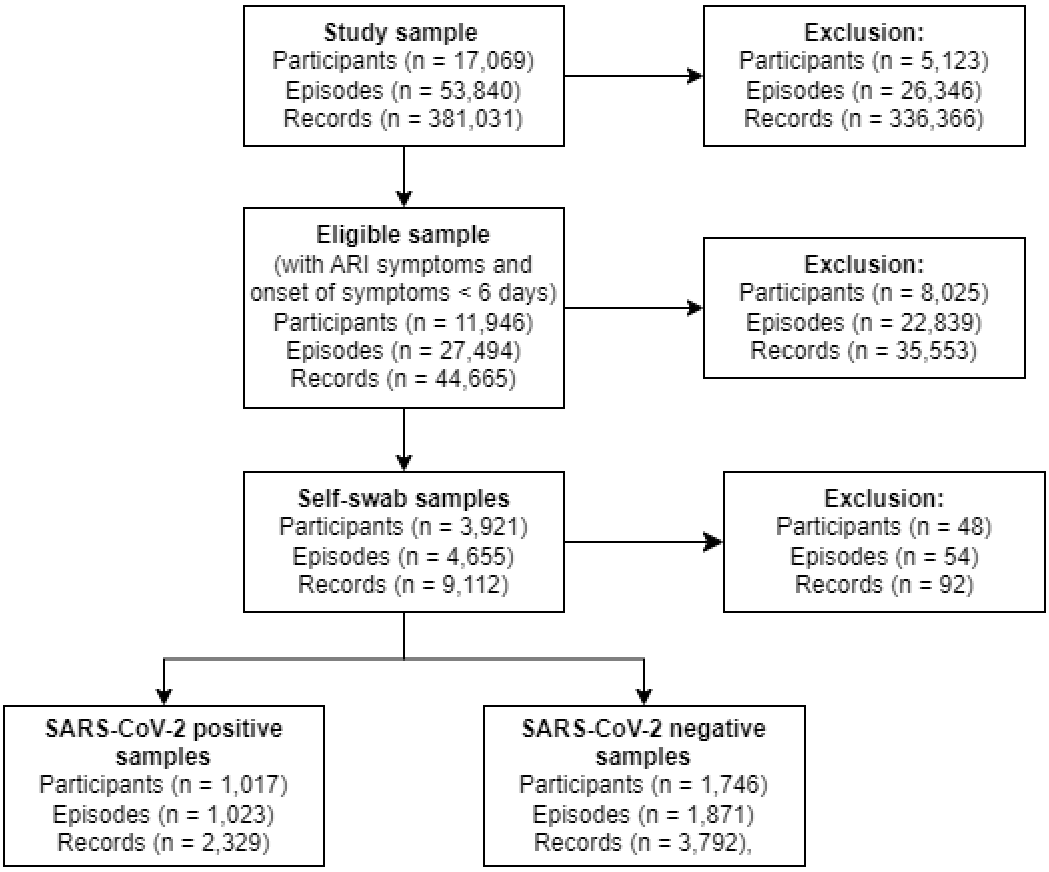


**Figure S1.** Flow diagram of the study population.

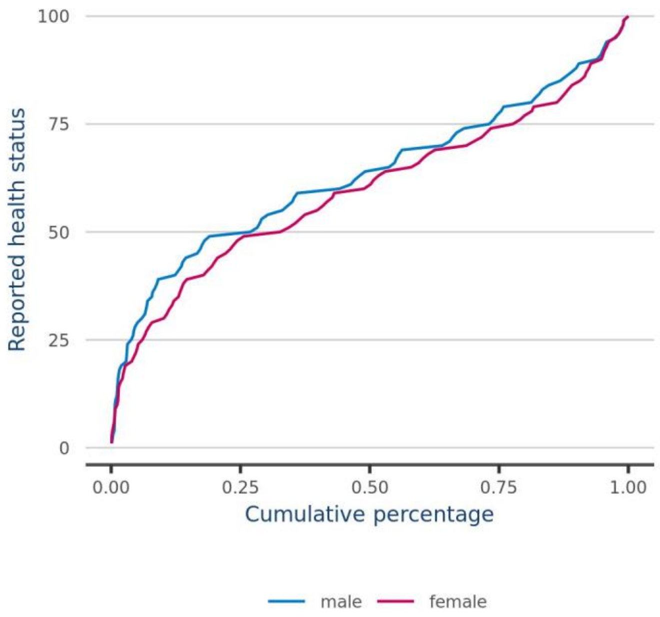


**Figure S2.** Distribution of health status among all participants with detected SARS-CoV-2 infection; males and females are plotted separately.

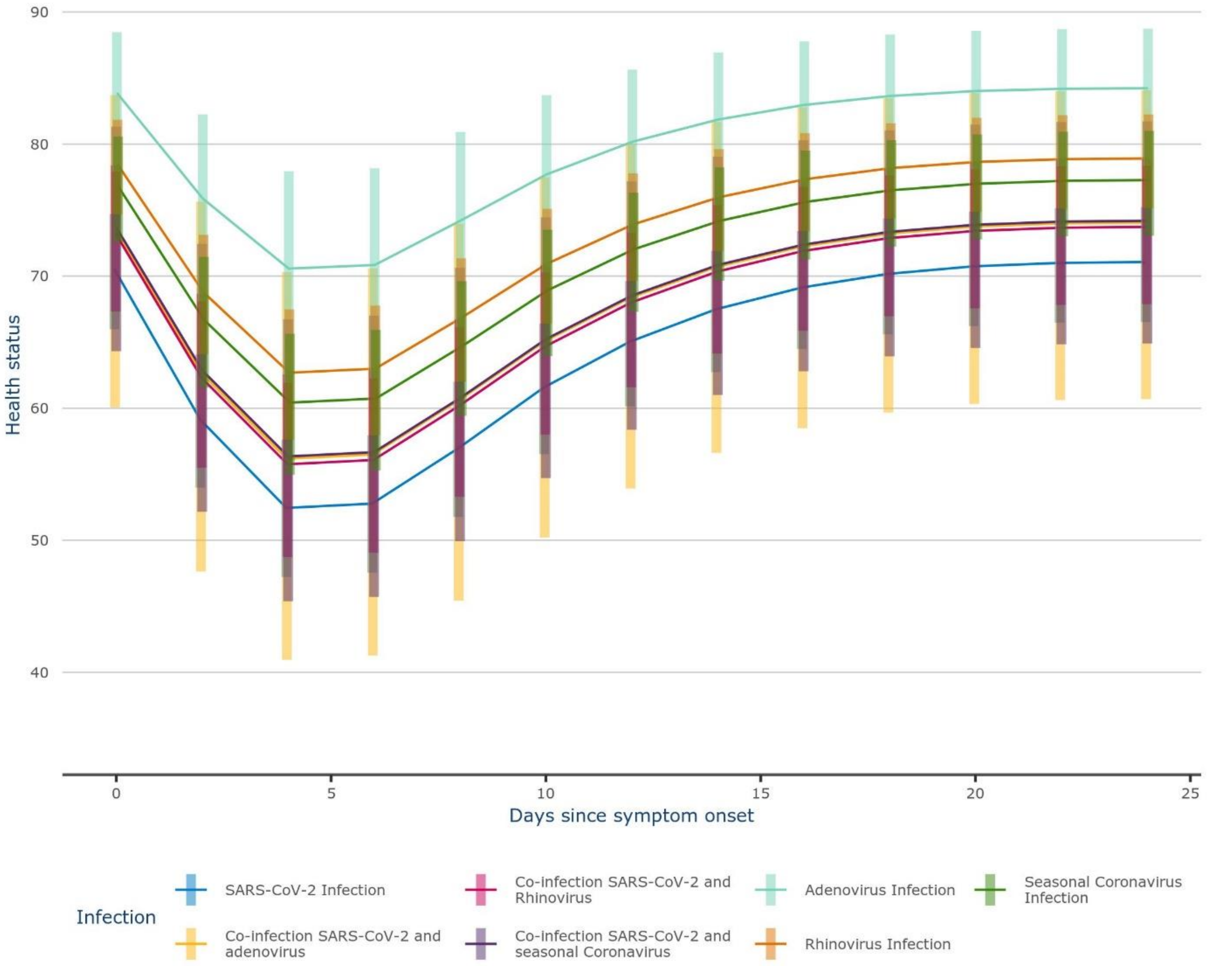


**Figure S3.** Estimated marginal means of the reported health status over time since symptom onset with 95% confidence level for participants with SARS-CoV-2, rhinovirus, seasonal coronaviruses and adenovirus mono-infection, and respective co-infection with SARS-CoV-2.

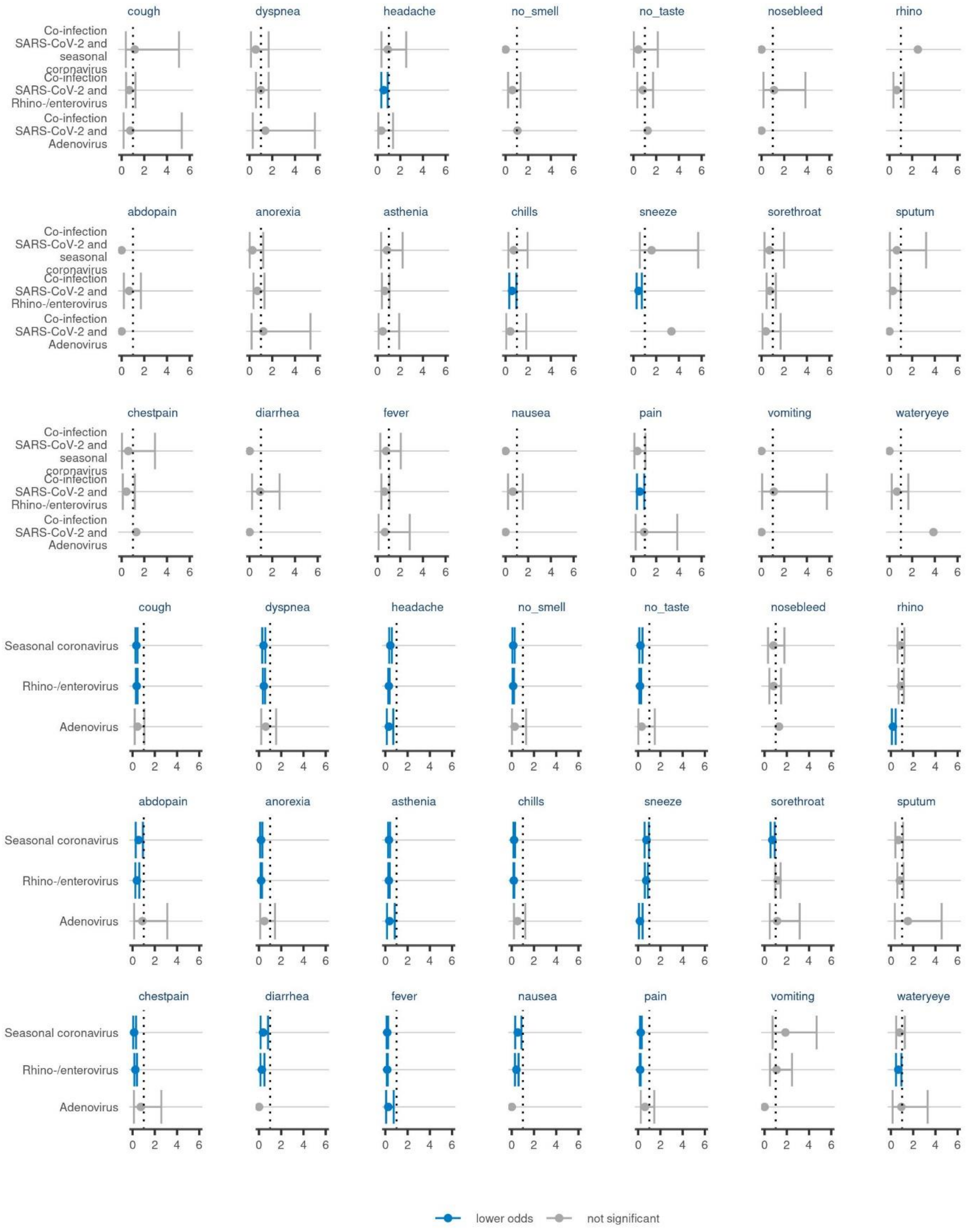


**Figure S4.** Adjusted odds ratio per symptom and infection compared to SARS-CoV-2 mono-infection. Adjusted for age group and sex.
